# COVID-19: A comparative study of severity of patients hospitalized during the first and the second wave in South Africa

**DOI:** 10.1101/2021.05.11.21257033

**Authors:** Caroline Maslo, Angeliki Messina, Anchen Laubscher, Mande Toubkin, Liza Sitharam, Charles Feldman, Guy A Richards

## Abstract

**Background:** South Africa has experienced two waves of COVID-19 infections, the second of which was inter alia attributed to the emergence of a novel SARS-CoV2 variant, 501Y.V2. This variant possibly has increased virulence and may be associated with increased mortality. The objective of this study was to determine if patients admitted in the second wave had more severe illness and higher mortality than those admitted in the first.

**Methods:** We analysed and compared the characteristics, biological severity markers, treatments, level of care and outcomes of patients hospitalised in a private hospital in the Eastern Cape Province, South Africa.

**Results:** Compared to the first wave, patients admitted in the second were older and less likely to have co-morbidities. In contrast, the D-dimer and interleukin-6 (IL-6) levels were significantly higher. Despite this, significantly less patients were admitted to ICU and/or were mechanically ventilated. The total length of hospital stay was identical in both groups. Whereas the overall mortality was not significantly higher during the second wave, the ICU mortality was. Those that died in the second wave were older than those in the first wave. Multivariable logistic regression showed that being admitted during the second wave was an independent risk factor for mortality.

**Conclusion:** This study appears to confirm previous reports that the 501Y.V2 variant is possibly more virulent as indicated by the higher levels of D-dimer and IL-6, the slight increase in mortality of hospitalised patients and the higher ICU mortality in the second wave.

## INTRODUCTION

The Severe Acute Respiratory Syndrome Coronavirus-2 (SARS-CoV-2) which causes COVID-19, has given rise to a global pandemic. Many countries have seen multiple waves of infection, with South Africa having experienced two [1]. South Africa reported its first case on 2 March 2020, with the epidemic curve peaking in the first week of July 2020, followed by a decline until the second week of September [2]. The second wave began as an isolated cluster of cases in October 2020 in the Eastern Cape Province (ECP) of South Africa and then spread rapidly to involve the rest of the country. By October 15^th^, all samples routinely genotyped in the ECP were positive for 501Y.V2 [3] and the rapidly accelerating incidence of cases in that province, and nationally, was putatively linked to the emergence of a novel SARS-CoV-2 variant, 501Y.V2 or B.1.351.

As new variants emerge, a select few have demonstrated increased transmissibility and possibly clinical severity [4-5]. Until November 2020, there had been a shift towards patients that were younger, with fewer associated comorbidities and lower in-hospital mortality. The latter was probably multifactorial and linked to the younger age, the lower comorbidity burden, possibly better medical management and less severe disease [1, 6-9]. However, the B 1.1.7 variant that emerged in November 2020 in the south of England and spread to both Europe and the United States (US) has been associated with higher transmissibility and mortality [4-5-10]. Recently, Pearson et al., using a model developed in the United Kingdom (UK) found evidence indicating that the 501Y.V2 variant may cause more severe disease [11]. Similarly, the South African National Institute for Communicable Diseases (NICD), by analysing the data from the National Active Surveillance system for COVID-19 hospitalisations, has reported a 20% increased mortality among individuals hospitalised with COVID-19 during the second wave [12].

In an attempt to confirm whether the 501Y.V2 variant does confer an increase in severity, we conducted a study to assess characteristics, severity of illness and mortality in patients hospitalised during the second wave and compared this with those hospitalised in the first, in a private hospital in the ECP.

## METHODS

### Study design

We conducted a retrospective study of hospitalised cases of SARS-CoV-2 infection in Greenacres Hospital (a member of the Netcare group of Hospitals) in the ECP, South Africa. The first COVID-19 patient was admitted on 28 March 2020, and thereafter the number of admissions increased to peak in the week of 14 June 2020 (week 14) and decreased until 22 August 2020. The second wave started on 25 October 2020 to peak on 8 November 2020 (week 34) and decreased from 6 December 2020 to 24 January 2021. We extracted data from two equal six-week periods in each of the waves, during both of which there were continuous admission rates of more than 40 patients per week (Fig 1). We categorized patients into the first wave for admissions between 14 June and 25 July 2020 (weeks 13 to 18) and the second wave for admissions between 8 November to 19 December 2020 (weeks 34 to 39). The criteria for enrolment in the study were hospitalisation and a positive reverse transcriptase polymerase chain reaction (RT-PCR) obtained from a nasopharyngeal swab in consecutive hospitalised patients. Patients not admitted for COVID-19 and those that died on admission were excluded from the study.

**Figure 1.**
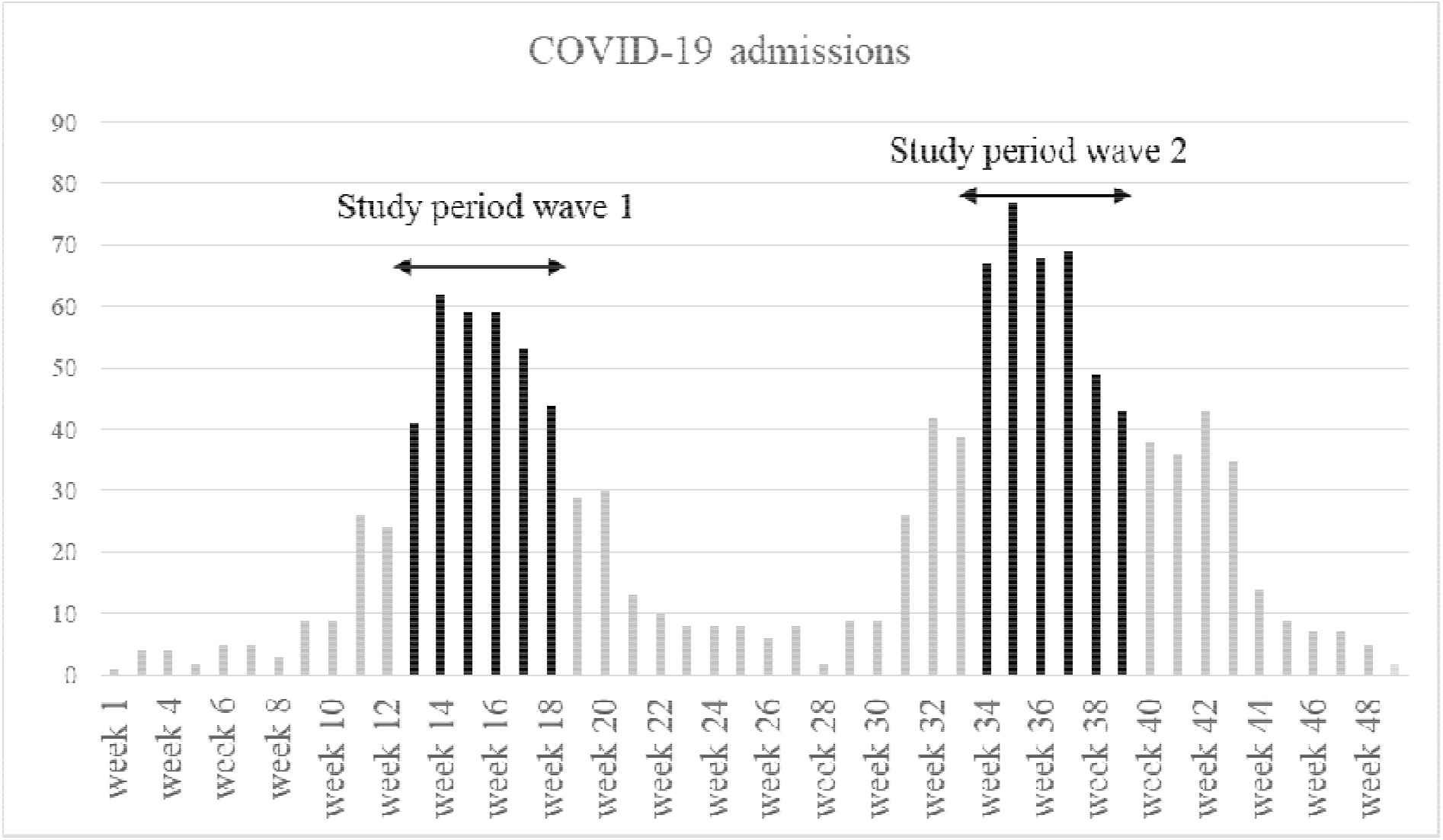
Weekly distribution of COVID-19 admissions during the first and second waves in the ECP, South Africa.

This study was approved by Pharma-ethics as complying with the Ethical Standards of Clinical Research based on current U.S. Food and Drug Administration (FDA), International Conference for Harmonization (ICH), Good Clinical Practice (GCP), Declaration of Helsinki and current local Ethics guidelines (reference 200923584).

### Data collected

Patient characteristics, therapy, highest level of care required, need for mechanical ventilation, length of stay and mortality were extracted from the Netcare electronic billing system (SAP ISH-SAP ERP ENHANCE package, version 6.07). Laboratory data were collected from the Bluebird Health platform (Intelligent Medical Systems, South Africa).

### Statistical analysis

Data is provided as numbers and percentages. Continuous variables are presented as medians and interquartile range (IQR) or means and standard deviations, as appropriate. Statistical comparisons between the two groups were made using the Mann-Whitney U test or the Student’s t test for continuous variables, with the use of the Fisher’s exact test for categorical variables. Analysis of factors potentially associated with mortality was performed using multivariate logistic regression. Statistical significance was set at p< 0.05. All statistical analyses were conducted using IBM SPSS version 27.0.1.0.

## RESULTS

Overall, 550 patients were admitted during the first wave and 685 patients during the second. The distribution of COVID-19 hospital admissions is shown in figure 1. During the designated study periods, 303 patients with COVID-19 were admitted during the first wave and 357 patients during the second.

The patients’ epidemiological characteristics are shown in table 1. Those admitted during the second wave were significantly older (median 57 vs 54 years; *p= 0*.*03*) and there were more patients without any comorbidities (47% vs 36.9%; *p= 0*.*008*). In addition, less patients had hypertension (32.2% vs 45.2%; *p< 0*.*001*) but there was no difference in the prevalence of diabetes mellitus, chronic cardiorespiratory or cerebrovascular diseases, or HIV positivity. The laboratory data on admission indicated that the D-dimer and interleukin-6 (IL-6) levels were higher in patients admitted during the second wave (1.1 vs 0.9 mg/l, *p= 0*.*005* and 55.1 vs 32.4 pg/ml, *p < 0*.*001*, respectively) while no difference was observed in the C-reactive protein (CRP) levels, or the neutrophil to lymphocyte ratio (NLR) (Table 2).

**Table 1.** Comparison of characteristics of patients admitted with COVID-19 in the first and second waves in the ECP, South Africa.

|  | First wave<br>(No=303) | Second wave<br>(No=357) | *P* value |
| --- | --- | --- | --- |
| **Patients characteristics** |  |  |  |
| Sex (Male), No (%) | 166 (54.7%) | 179 (50.1%) | 0.23 |
| Age (y), median (IQR) | 54 (15.4) | 57 (20) | 0.03 |
| Age at death (y), median (IQR) | 60 (15) | 65 (20) | 0.008 |
| **Age categories** | No (%) | No (%) |  |
| 0-29 years | 8(3%) | 14 (4%) | 0.488 |
| 30-59 years | 193 (64%) | 182 (51%) | <0.001 |
| ≥60 years | 102 (34%) | 161 (45%) | 0.004 |
| **Main co-morbidities** | No (%) | No (%) |  |
| Diabetes mellitus | 82 (27.0%) | 99 (27.7%) | 0.84 |
| Hypertension | 137 (45.2%) | 115 (32.2%) | 0.0006 |
| Chronic respiratory disease | 9 (3%) | 16 (4.5%) | 0.31 |
| HIV infection | 6 (2%) | 4 (1.1%) | 0.34 |
| Cardiovascular disease | 60 (16.5%) | 78 (21.8%) | 0.08 |
| Cerebrovascular disease | 1 (0.3%) | 5 (1.4%) | 0.13 |
| No co-morbidities | 112 (36.9%) | 168 (47.0%) | 0.008 |
| No., number; IQR, interquartile range; HIV, human immunodeficiency virus |  |  |  |

**Table 2.** Comparison of severity markers levels at admission of patients between the first and second waves.

|  | Normal | First wave | Second wave | *P* value |
| --- | --- | --- | --- | --- |
|  | range | Median (IQR) | Median (IQR) |  |
| D-dimers (mg/L) | 0-0.5 | 0.9 (1.01) | 1.1 (1.46) | 0.005 |
| N/L ratio | 1-3 | 5 (6) | 6 (6) | 0.27 |
| C-reactive protein<br>(mg/L) | 0-10 | 134 (1.15) | 135 (1.005) | 0.61 |
| Interleukin-6 (pg/ml) | < 6.4 | 32.4 (64.5) | 55.1 (96.2) | <0.001 |
| Hs-Troponin (ng/L) | 0-70 | 14.7 [21.6] | 13.8 [37.5] | 0.99 |
| N/L ratio: Neutrophil-to Lymphocytes ratio |  |  |  |  |

Table 3 summarises medications and supportive treatment. Over 90% of patients in both waves received glucocorticoid and anticoagulant therapy. Remdesivir was not available in South Africa during the first wave study period, but was prescribed in 37.2% of patients in the second. There was also an increase in prescription of tocilizumab in the second wave as compared to the first (49.5% vs 20.8%, *p= 0*.*03*). Less patients were admitted to the intensive care unit (ICU) in the second wave versus the first (35% vs 48.5%; *p= 0*.*03*) and fewer patients received mechanical ventilation during the second wave as compared to the first (8.9% vs 15.5%, *p= 0*.*009*).

**Table 3.** Comparison of treatment administered in the first and second waves in the ECP, South Africa

|  | No. (%) |  | *p*-value |
| --- | --- | --- | --- |
|  | First wave (n=303) | Second wave (n=357) |  |
| Glucocorticoids | 273 (90.0%) | 323 (90.4%) | 0.86 |
| Thromboprophylaxis | 276 (91.0%) | 331 (92.7%) | 0.42 |
| Remdesivir | Not available | 133 (37.2%) | N.A |
| Tocilizumab | 63 (20.8%) | 177 (49.5%) | <0.001 |
| Number of ICU admissions | 147 (48.5%) | 125 (35%) | <0.001 |
| Invasive mechanical ventilation | 47 (15.5%) | 32 (8.9%) | 0.009 |
| N.A not applicable; No.: number |  |  |  |

As shown in table 4, the number of days between hospital and ICU admission was also longer in the second wave (3.13 days vs 1.55 days, *p= 0*.*01*) and fewer patients were admitted to the ICU within 24 hours of hospital admission. There was no difference in hospital and the ICU length of stay between the two waves. Overall mortality was 32.6% in the first wave and 36.4% in the second. However, ICU mortality was significantly higher in the second compared with the first (74.4% vs 57.1%, *p= 0*.*002*).

**Table 4.** Main outcomes of patients hospitalized during the first and second waves in the ECP, South Africa

| | Mean ± SD | | *p*-value |
| --- | --- | --- | --- |
|  | First wave (n=303) | Second wave (n=357) |  |
| Days between hospital admission and death | 11.74 ± 7.63 | 11.14 ± 9.26 | 0.6 |
| Days between ICU admission and death | 10.67 ± 7.58 | 10.32 ± 8.84 | 0.72 |
| Length of ICU stay, days | 11.24 ± 11.16 | 9.20 ± 9.11 | 0.1 |
| Length of hospital stay, days | 10.49 ± 9.8 | 10.20 ± 8.01 | 0.67 |
| Days between hospital and ICU admission | 1.55 ± 2.32 | 3.13 ± 5.46 | 0.001 |
| Outcomes | No.(%) |  | *p*-value |
|  | First wave (n=303) | Second wave (n=357) |  |
| Admission to ICU ≤ 24 hours | 97/147 (65.9%) | 63/125 (50.4%) | 0.01 |
| Secondary infections | 5 (1.6%) | 5 (1.4%) | 0.83 |
| Intensive care unit mortality | 84/147 (57.1%) | 93/125 (74.4%) | 0.002 |
| Overall mortality | 98/303 (32.3%) | 130/357 (36.4%) | 0.26 |

SD: Standard deviation; No.: number

Multivariate analysis of factors associated with mortality identified age, ICU admission, mechanical ventilation, treatment with tocilizumab and admission during the second wave as independently associated with death (Table 5).

**Table 5.** Multiple regression model investigating risk factors for death in the ECP, South Africa

|  | **B** | **SE** | **Exp(B)** | ***p*-value** |
| --- | --- | --- | --- | --- |
| Age | 0.910 | 0.631 | 1.095 | <0.001 |
| Gender | -0.129 | 0.231 | 0.879 | 0.577 |
| Absence of comorbidity | 0.410 | 0.237 | 1.507 | 0.083 |
| ICU admission | -2.804 | 0.292 | 0.061 | <0.001 |
| Use of invasive mechanical ventilation | 2.011 | 0.398 | 7.47 | <0.001 |
| Remdesivir | -0.563 | 0.318 | 0.57 | 0.07 |
| Tocilizumab | 0.677 | 0.256 | 1.968 | 0.008 |
| Admission during the second wave | 0.789 | 0.287 | 2.202 | 0.006 |

B: Non-standardized $\beta$ coefficient. SE: Standard error of B.

## DISCUSSION

In contrast with most reports of second waves outside of South Africa, our study found that patients hospitalised during the second wave in a single hospital in the ECP were significantly older than in the first [6-8]. Of note, the second wave coincided with the end of the school and university years and also the annual summer public holidays in which there was significant liberalisation of lockdown rules. As such, spread among younger people would have been expected to be greater.

Patients hospitalised during the second wave had fewer co-morbidities, which was noted in some studies but was usually associated with younger age of the patients. One plausible explanation would be related to behavioural change in those with chronic conditions as they became more aware of an increased risk for adverse outcomes.

In the current study, laboratory parameters indicating increased risk of severe COVID-19 such as N/L ratio and CRP were identical in both groups, but D-dimer and IL-6 levels were higher on admission in the second wave. It has previously been suggested by Wungu et al., in a meta-analysis comparing cardiac markers as predictive factors in COVID-19 infection that the D-dimer level was the best predictor of severity and mortality [13]. IL-6 levels were also shown to be significantly elevated in patients with severe disease relative to those with less severe disease. In a recent study from Sabaka et al., among patients with COVID-19 in a long-term care facility, IL-6 was the most robust predictor of hypoxemia when compared to other laboratory parameters such as the CRP and D-dimer levels [14]. The higher levels of IL-6 and D-dimer levels in the current study do, therefore, suggest that those admitted in the second period had higher disease acuity on admission.

In contrast, the proportion of patients admitted to the ICU and the proportion that received mechanical ventilation were lower in the second wave. ICU admissions within the first 24 hours of following hospital admission were also lower in the second wave. The latter has been a worldwide phenomenon, and is related to the fact that it is currently recognised that respiratory failure could be effectively treated with high flow nasal oxygen treatment without the necessity for early intubation [15]. While lack of capacity in the ICU setting may have contributed to the higher mortality, the lower absolute number of patients admitted to the ICU in the second wave suggest that shortage of ICU beds was not an issue in the current study.

Prescription of corticosteroids and anticoagulants were similar in both groups. Remdesivir was not available during the first study period but was prescribed to more than a third of patients during the second. A greater proportion of patients in the second wave also received tocilizumab as doctors became more familiar with its use and possibly also because patients in the second wave had higher IL-6 levels. Both remdesivir and tocilizumab were initially reported to be effective in shortening length of stay and preventing death [16, 17]; however, this remains controversial [18, 19].

Whereas it could be assumed that the knowledge and experience gained from managing patients in the first wave would have impacted positively on the outcomes in the second wave, we observed a significant increase in mortality and shorter time to death (albeit not significant) in those admitted to ICU during this period.

Other studies have reported a similar lack of improvement in mortality in patients admitted to ICU between the first and the second waves despite apparently better management. Karagiannidis *et al*., reported a 50% drop in ICU admissions during the second wave of the pandemic in Germany but in contrast the prognosis of ICU patients, remained unchanged, with the mortality greater than 50% [20]. Contou *et al*., reported their experience of ICU admissions during the second wave in France and did not observe any decrease in ICU mortality which remained at 50% [21]. Of note, none of those patients received remdesivir or tocilizumab.

It is probable that patients that were transferred to ICU were more severe in the second wave than in the first, as both the IL-6 and D-dimer levels were elevated to a greater extent than in the first, and also because patients were managed for longer in the general wards on non-invasive respiratory support prior to ICU admission. This potentially could mean that those that failed to respond to these oxygen delivery devices were those with progressive disease who consequently had a worse prognosis.

Finally, the multivariate regression analysis identified admission during the second wave as an independent factor associated with death. Patients in the second wave were each assumed to be infected with the 501Y.V2 lineage of SARS-CoV2 as, since as mentioned above, all samples that had been routinely genotyped in the Eastern Cape Province were positive for 501Y.V2 from October 2020 [3]. For all these reasons, the current study potentially confirms the increased severity of this variant as had been previously reported in the National Surveillance study [12].

There are limitations in our study. This was a single centre study and thus the results cannot be generalised to other regions in South Africa, nor to other countries. Patient numbers were, however, adequate. Furthermore, being a single centre study, it also excluded our need to have to evaluate the impact that different skill sets and resource availability at different hospitals may have had on the outcomes. We could not perform genotyping for all patients included in the study and thus relied on screening results performed by the Kwazulu-Natal Research Innovation and Sequencing Platform (KRISP), which indicated that all the patients genotyped in the second wave in the ECP had the variant. As most data were extracted from our billing platform, certain clinical data were missing. Obesity, which is known to be an important risk factor for COVID-19 severity, was not evaluated as the body mass indices were not recorded [22]. Data on alternative non-invasive oxygen strategies, such as high frequency nasal oxygen delivery devices were also not available. Patients in the second wave did have higher IL-6 and D-dimer levels on admission but the date of symptom onset was not recorded and although admission delays could not be excluded, all patients presenting via the emergency department were admitted promptly.

## CONCLUSION

This study indicates that the IL-6 and D-dimer levels were higher in patients during the second wave compared with the first, the ICU mortality was significantly higher, and the multivariate analysis showed that admission within the second wave was independently associated with mortality. As such it is possible that 501Y.V2 variant is more virulent than the original variant although confounders are present.

## Data Availability

The dataset used and analysed for the current study is available from the corresponding author on reasonable request.

## Notes

### Author’s contribution

CM: conception, methodology, formal analysis, writing original draft, writing-review and editing. AL: supervision, resources and writing-review. MT and LS: data acquisition. AM: medication data and writing-review. CF and GR: review and interpretation of results, writing-review and editing. All authors read, critically reviewed and approved the final version.

## Acknowledgments

The authors warmly acknowledge Dr M. Butae, Dr J. Dryer, Dr N. Hendricks, Dr. L Kruger, Dr M. Jansen, Dr Y. Jansen, Dr H.Maepa, Dr S. Meel, Dr P. Meel, Dr P. Musoke, Dr D. Porter, Dr H. Potgieter and all the staff of Greenacres Hospital who have tirelessly provided care for COVID-19 patients during this pandemic.

## Ethics approval

This study was approved by Pharma-ethics as complying with the Ethics Standard of Clinical research based on current FDA, ICH, GCP, Declaration of Helsinki and current local Ethics under the number 200923584.

